# Educational Inequalities in Well-Being in Later Life in Germany: The Role of Health Behaviours and Health Literacy

**DOI:** 10.64898/2026.04.22.26351388

**Authors:** Fabio Franzese, Michael Bergmann, Agnieszka Burzynska

## Abstract

Socioeconomic inequalities in health and well-being are a major public health concern, particularly in ageing populations. Education is a key determinant shaping multiple aspects of health outcomes. We used cross-sectional data from wave 9 of the German sample (n=4,148) of the Survey of Health, Ageing and Retirement in Europe (SHARE) to test whether formal education is associated with well-being in later adulthood, with health literacy, self-rated health, and preventive health behaviours as possible mediators. Our results showed that education was positively associated with greater well-being, but only via indirect pathways. Specifically, self-rated health, health literacy, and fruit and vegetable consumption mediated the relationship between education and well-being accounting for 54.7, 24.7, and 12.6 percent of the total effect, respectively. In addition, there were significant positive correlations between education and health literacy, as well as high-intensity physical activity, daily fruit and vegetable consumption, more preventive health check-ups, and less smoking. In contrast, alcohol consumption was more common among those with higher levels of education. All health behaviours and health literacy were correlated directly or indirectly (i.e., mediated by health) with well-being. These findings highlight the importance of examining indirect pathways linking education to well-being in later life. Interventions aimed at improving health literacy and promoting healthy behaviours may help reduce educational inequalities in quality of life among older adults.

**About the SHARE Working Paper Series:** The SHARE Working Paper Series started in 2011 and collects pre-publication versions of papers or book chapters, technical and methodological reports as well as policy papers based on SHARE data. The working papers are not reviewed by the publisher (SHARE-ERIC), layout and editing are not standardized. The publisher takes no responsibility for the scientific content of the paper. Working Papers can be updated – a version number is indicated on the front page. Previous versions are available upon request.

## 1 Background

Education has long been recognised as a key social determinant influencing various aspects of life, including health, health-behaviours, and overall well-being. While formal education demonstrably structures life chances and trajectories early in life, e.g. in the job market (Hillmert, 2017), its enduring role beyond midlife is increasingly acknowledged. However, the underlying mechanisms and pathways through which education continues to influence well-being are not yet fully understood.

Health behaviours encompass all actions and habits that influence health, such as physical activity, healthy diet, and participation in preventive health check-ups. Commonly, higher educational attainment is associated with more favourable health behaviours (A. Richter et al., 2021). However, empirical evidence suggests that this relationship may vary depending on individual characteristics, such as age, sex, and socio-economic status (SES) as well as contextual factors, such as cultural and regional settings. For instance, in the German population, individuals with higher education levels tend to engage more in leisure-time physical activity. Paradoxically, they also report higher levels of sedentary behaviour, primarily attributed to a greater proportion of sedentary workplaces (Manz et al., 2022; Nowossadeck and Spuling, 2025). Furthermore, a counter-intuitive pattern emerges regarding alcohol consumption, with higher rates of risky alcohol consumption often observed among highly educated individuals in the general German population (Lange et al., 2017; A. Richter et al., 2025). Regarding preventive health checks in Germany, a positive association has been found with a composite index of socio-economic status (SES, including income, education, and occupation), but not consistently with education as a single indicator (Hoebel et al., 2013; M. Richter et al., 2002). These findings highlight the complex and sometimes contradictory nature of the relationship between education and various health behaviours.

Health literacy is the capacity to access, understand, and use health information to make informed decisions. It represents a critical resource, especially in older age, as individuals face increasing interactions with complex healthcare situations and treatment decisions. Consistent findings demonstrate a strong positive correlation between educational attainment and health literacy (Kolpatzik et al., 2025). Health literacy can also foster self-efficacy (Lee, 2016) and improve medical decision-making (Seo, 2016), thereby potentially influencing health outcomes and well-being.

Education is also a well-documented correlate of well-being in old age (Alcañiz and Solé-Auró, 2018; Mendorf et al., 2023; Powdthavee et al., 2015; Svendsen et al., 2020). While various concepts and definitions of well-being exist, health-related quality of life is often considered particularly relevant in older age. Previous research has identified health and income as main pathways through which education can influence life satisfaction and health-related quality of life (Powdthavee et al., 2015; Sheikh et al., 2017). However, tests of other potential mediators beyond traditional socio-economic indicators are scarce.

Preventive health behaviours, such as physical activity, healthy nutrition, and avoidance of harmful substances, are central pathways linking education to health and well-being (Laaksonen et al., 2008; Moor et al., 2017; Petrovic et al., 2018). Physical activity, for example, has been reported as a significant predictor for quality of life in old age (Lima et al., 2011; Mendorf et al., 2023), suggesting its potential mediating role in the education-well-being relationship.

Furthermore, health literacy is a well-known mediator between socio-economic status and health outcomes (Stormacq et al., 2019), and its direct influence on wellbeing, potentially by improving navigation within the healthcare system, enhancing coping mechanisms for illness, and promoting active lifestyle choices, warrants further investigation. Despite the recognised importance of education, health behaviours, and health literacy, empirical evidence on their interrelations and their ultimate impact on well-being among older adults remains limited. Many existing studies utilize composite SES scores rather than examining the unique role of education. Furthermore, studies investigating mediation effects, such as education leading to lifestyle behaviours and then to health, often employ composite lifestyle behaviour scores and rarely focus specifically on older populations. This gap in the literature underscores the need for a more nuanced understanding of these relationships.

This study aims to investigate the interrelations between formal education, health literacy, specific preventive health behaviours, and well-being in later adulthood in Germany. More specifically, we address the following research questions:

1. Is formal education associated with health literacy and preventive health behaviours (physical activity, alcohol consumption, smoking, nutrition, preventive check-ups) in later life in Germany?
2. Do health literacy and specific preventive health behaviours mediate the relationship between formal education and well-being in later life?

## 2 Data and Method

### 2.1 Sample

The analysis of associations between education, health literacy, health behaviour, and well-being used cross-sectional data from Wave 9 of the German data collection of the Survey of Health, Ageing and Retirement in Europe (SHARE) (SHARE-ERIC, 2024b). SHARE is an interdisciplinary panel study of the population aged 50 and older conducted via face-to-face interviews (Börsch-Supan et al., 2013).

A total of 4,467 respondents is included in the German SHARE Wave 9 data. Thirty-two individuals were excluded as they were younger than 50 at the time of interview (younger spouses that were interviewed as partners). Cases with missing information in one of the variables used for analysis were excluded. Most missing information were present in the indicators for quality of life (184 cases), memory test (100 cases), and financial situation – making ends meet (61 cases). Only the household income – that was used to calculate an indicator for poverty – was imputed (using the imputations provided by SHARE) because the share of missing information was higher than 10 percent. After exclusion of these cases, a total of 4,148 individuals were available for analysis.

### 2.2 Measures

We used the level of education attained based on the Internal Standard Classification of Education 1997 (ISCED-97) as the independent variable. Respondents were grouped into three categories (Avendano et al., 2009): primary education (ISCED-97 score: 0–2), secondary education (ISCED-97 score: 3), and post-secondary education (ISCED-97 score: 4–6).

Well-being, the dependent variable, was measured using the CASP-12 scale that measures quality of life of older adults and comprises four dimensions: control, autonomy, self-realisation, and pleasure (Hyde et al., 2003). Each dimension was evaluated by three questions on a four-point scale. The total score for all twelve questions ranged from 12 to 48, with higher values representing a better quality of life.

Health literacy was assessed using the single item literacy screener (SILS) (Morris et al., 2006). SILS is an instrument that captures the need for support with written health information, e.g. from doctors or pharmacy, by asking “How often do you need to have someone help you when you read instructions, pamphlets, or other written material from your doctor or pharmacy?”. Answer options are “always”, “often”, “sometimes”, “rarely”, and “never”. A binary indicator for high reading related health literacy (“never”) is used in the analysis.

The study included five self-reported indicators of health behaviours. High physical activity was defined as engaging in vigorous physical activity more than once a week, where vigorous activity was described in the questionnaire as “such as sports, heavy housework, or a job that involves physical labour”. A healthy nutrition indicator was defined as daily consumption of fruit or vegetables. Alcohol consumption was defined as having one or more alcoholic drinks in the last 7 days. Smoking behaviour was defined as respondents identifying themselves as “currently smoking”. High physical activity, daily fruit and vegetable consumption, alcohol consumption, and smoking were coded as binary indicators. A preventive healthcare utilisation score was constructed by summing scores on following items: 1) Having visited a dentist (including for a routine check-up) within the last 12 months; 2) received a flu vaccination within the last 12 months; 3) Having undergone colon cancer screening within the last two years; 4) Having had an eye examination within the last two years. Affirmative responses were coded as 1 and negative responses as 0, resulting in a total score ranging from 0 to 4.

Self-reported overall health was assessed with answer options “excellent”, “very good”, “good”, “fair”, “poor”, recoded to values 0 (poor) to 4 (excellent).

Confounding variables included age groups (50-64; 65-79; 80+), gender, presence of a partner in the same household, migration history (born in Germany vs. abroad), and good eyesight for reading. Financial situation was controlled for using a question on how the households was able to make ends meet (no difficulties, some difficulties, or great difficulties) and the household’s poverty risk, defined as having an equivalised disposable income of less than 60% of the median in the sample. Missing information on the household income was imputed based on the multiple imputations provided by SHARE (SHARE-ERIC, 2024a).

Cognitive function was assessed using the 10-word memory test (Brandt et al., 1988). Respondents were asked to immediately repeat ten words that were read out by the interviewer. A few minutes later, participants were asked to repeat all words they remember. In the analysis, the average of the immediate and delayed recall test was used as measure of cognitive function.

### 2.3 Statistical Analyses

Descriptive analyses were performed to examine distributions of health literary, health behaviours, and well-being across age groups and education levels. To ensure representativeness of the population aged 50 years and older in Germany, all analyses applied the provided calibration weights (SHARE-ERIC, 2024a).

To investigate potential mediation mechanisms, we performed path analyses using multiple linear regression models in Stata (version 17). In the path model, health, health literacy and health behaviours are considered as potential mediators for the association between education and well-being, see Figure 1. Indirect effects were estimated using a bootstrapping approach with 5000 repetitions to obtain robust confidence intervals, and Monte Carlo estimates were reported. This approach allows for a more reliable assessment of mediation effects, particularly in complex models.

**Figure 1.**
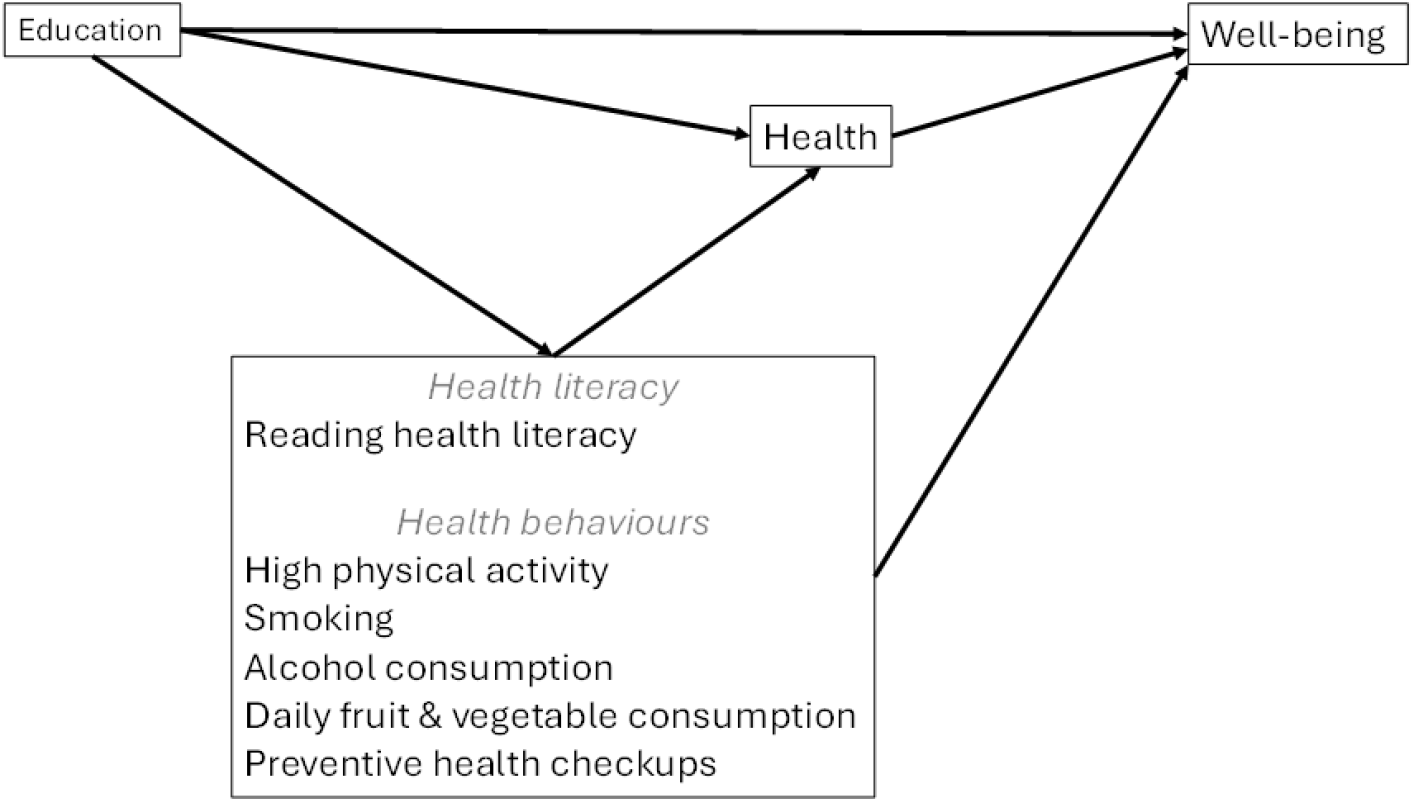
Path model for investigating the associations between education and well-being, with health, health literacy and health behaviours as possible mediators.

The results of the mediation analyses were interpreted following the framework proposed by Zhao et al., 2010, which distinguishes different types of mediation based on the presence, significance, and direction of direct and indirect effects. Specifically, *complementary mediation* is assumed when both direct and indirect effects are statistically significant and point in the same direction, whereas *competitive mediation* occurs when both effects are significant but operate in opposite directions. *Indirect-only mediation* is identified when a significant indirect effect is present in the absence of a direct effect, while *direct-only non-mediation* refers to cases where only the direct effect is significant. Finally, *no-effect non-mediation* is concluded when neither direct nor indirect effects reach statistical significance.

## 3 Results

### 3.1 Sample Description

As shown in Table 1, a total of 4,148 participants were included in the analyses, with an average age of 67.8 years (range: 50–96). Of these, 53.7% were female and 46.3% were male. Approximately half of the sample (49.3%) had medium levels of education, 42.8% had high levels of education, and only 7.9% had low levels of education. The average CASP score was 39.1. In terms of health-related indicators and lifestyle, participants reported an average self-rated health score of 2.8 (on a 5-point scale ranging from 0 to 4, with higher values indicating better health). A high proportion (84.3%) demonstrated high reading health literacy. Physical activity levels showed that 40.9% of the sample reported engaging in high physical activity at least once per week, while 74.0% consumed fruit and vegetables daily. Alcohol consumption in the last 7 days was reported by 62.5% of the participants, and 16.1% were current smokers. The average number of preventive health check-ups was 2.3 out of 4. Social and cognitive aspects revealed that a significant majority (72.9%) of participants lived with a partner in the same household. A small proportion (10.1%) were born outside Germany. On the 10-word memory test, the average score was 5.1. Visual health was generally good, with 90.1% reporting good eyesight for reading. Financial well-being indicators showed that 2.6% of participants experienced great difficulties making ends meet, and 8.3% reported some difficulties, while the remainder reported no difficulties. Overall, 13.1% were identified as being at risk of poverty, and 11.6% of the sample had their income imputed for this assessment.

**Table 1:** Sample description.

| Variable | Statistic | Value |
| --- | --- | --- |
| Age | mean | 67.8 |
| Age: 50-64 | % | 41.1 |
| Age: 65-79 | % | 44.8 |
| Age: 80+ | % | 14.1 |
| Male | % | 46.3 |
| Female | % | 53.7 |
| Low education | % | 7.9 |
| Medium education | % | 49.3 |
| High education | % | 42.8 |
| CASP (Quality of life) | mean | 39.1 |
| Self-rated health | mean | 2.8 |
| High reading health literacy | % | 84.3 |
| High physical activity | % | 40.9 |
| Daily fruit & vegetable consumption | % | 74.0 |
| Alcohol consumption | % | 62.5 |
| Current smoker | % | 16.1 |
| Preventive health check-ups | mean | 2.3 |
| Partner lives in same household | % | 72.9 |
| Born outside Germany | % | 10.1 |
| 10-word memory test | mean | 5.1 |
| Good eyesight: reading | % | 90.1 |
| Make ends meet: great difficulties | % | 2.6 |
| Make ends meet: some difficulties | % | 8.3 |
| Poverty risk: Yes | % | 13.1 |
| Poverty risk: Income imputed | % | 11.6 |
| Total | N | 4,148 |

### 3.2 Health Literacy and Health Behaviours

Health literacy and health behaviour by age group and education is presented in Figure 2 (numbers in Appendix 1). Health-related outcomes show strong variation by age and, to a lesser extent, by level of education. Health literacy was highest among those aged 50-64 with high education, and lowest among those aged 80+ with low education, highlighting substantial disparities, particularly in the oldest age group. There were also differences in levels of physical activity by age, with the 80+ age group being the least active. Significant differences in activity levels by educational attainment can be observed in the 65-79 age group: those with a high level of educational attainment are the most active, while those with a low level of educational attainment are the least active.

**Figure 2.**
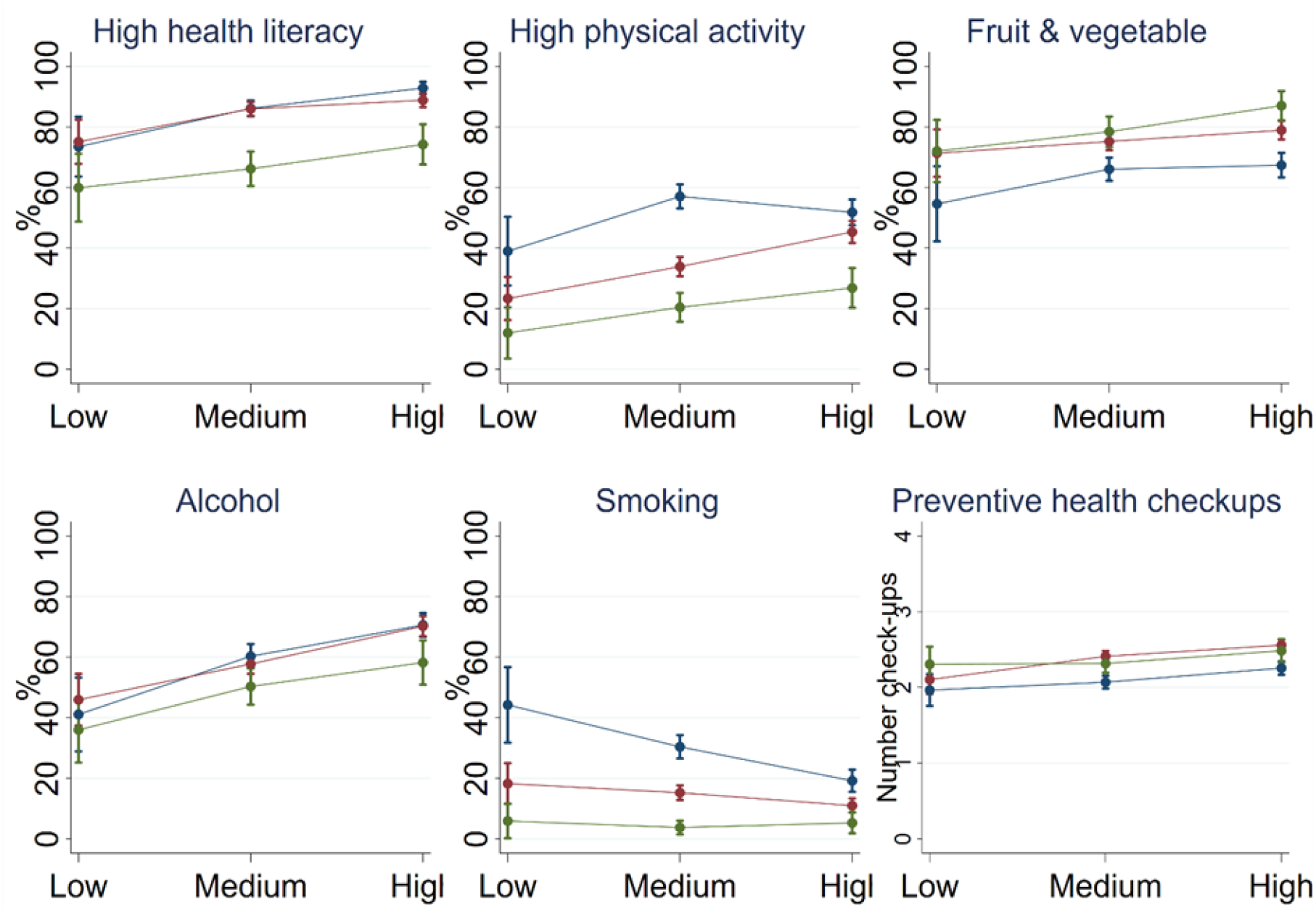
Health literacy and health behaviours by education and age groups. Data: SHARE Wave 9, Germany, weighted. N = 4,148.

Although there were no age-related differences within the low-educated group, older people in the highly educated group were more likely to consume fruit and vegetables daily (with significant differences observed across all age groups). The graph indicates a trend in educational attainment across age groups, although not significant.

Among 50-64-year-olds, a gradient in education was evident, with higher education being correlated with a greater likelihood of having consumed alcohol in the last 7 days (low education: 41.1% [95% CI: 28.9-53.3]; medium education: 60.3% [95% CI: 56.3-64.3]; high education 70.6% [95% CI: 66.7-74.6]). Similar differences in alcohol consumption by education were evident in other age groups: among 65-79-year-olds, for example, those with a higher education were more likely to consume alcohol than those with a medium or low education. Among those aged 80+, the high educated were more likely to consume alcohol than those with a low education. Older individuals seem to consume less alcohol, but age differences were only significant for those aged 80+ with a high level of education compared to younger adults.

A clear pattern by age emerged for smoking, with younger age groups being more likely to smoke, except among the highly educated, where the 65-79 age group is not significantly different from the 80+ age group. Among 50-64-year-olds, there were educational differences: the highly educated were significantly less likely to smoke than those with a medium or low education. Among those aged 80+, no significant differences by education were found.

The average number of preventive check-ups was similar across age and education. However, some significant differences were present. Highly educated individuals below 80 reported more check-ups than those with a medium level of education. Within the medium and high education groups, there were differences by age, with 65-79-year-olds having a higher number of checkups than the youngest age group.

Overall, these numbers indicate more pronounced differences by age than by education in the descriptive bivariate analyses. However, when examining the coefficients from the multivariate linear models (see Table 2, based on Appendix 3), a more nuanced picture emerged regarding the association of education with health-related indicators, after controlling for other factors. It is evident that education was significantly correlated with health literacy and all health behaviours even when various socio-demographic and economic variables were accounted for. Specifically, the multivariate models show that higher education was positively and significantly associated with a higher likelihood of high health literacy, greater physical activity, more frequent daily fruit and vegetable consumption, and increased alcohol consumption. Conversely, higher education was significantly associated with a lower likelihood of smoking. Furthermore, a strong positive association was present between education and the number of preventive health check-ups, suggesting that highly educated individuals tend to utilize preventive healthcare services more frequently. These multivariate findings underscore the independent and pervasive role of education as a crucial determinant of health and health-related behaviours, even when age-related variations are present.

**Table 2:**
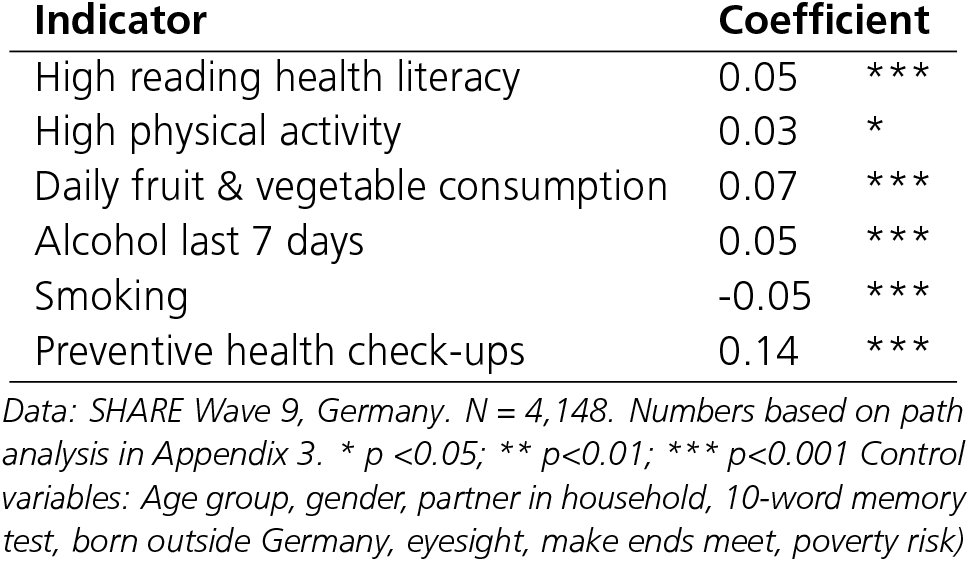
Correlations of health literacy and health behaviours with education (linear models, with control variables)

| Indicator | Coefficient |  |
| --- | --- | --- |
| High reading health literacy | 0.05 | *** |
| High physical activity | 0.03 | * |
| Daily fruit & vegetable consumption | 0.07 | *** |
| Alcohol last 7 days | 0.05 | *** |
| Smoking | -0.05 | *** |
| Preventive health check-ups | 0.14 | *** |

Data: SHARE Wave 9, Germany. $N = 4,148$ . Numbers based on path analysis in Appendix 3. \* $p < 0.05$ ; \*\* $p < 0.01$ ; \*\*\* $p < 0.001$ Control variables: Age group, gender, partner in household, 10-word memory test, born outside Germany, eyesight, make ends meet, poverty risk)

### 3.3 Education and Well-Being

As depicted in Figure 3 (see Appendix 2 for more detailed numbers), CASP index scores for quality of life consistently varied by educational attainment across all observed age groups. Among 50-64-year-olds, participants with high education reported significantly higher CASP scores than those with either low or medium education. A distinct educational gradient in quality of life was also apparent in the 65-79 age group, where the highly educated exhibited the highest scores and the least educated the lowest. Among individuals aged 80 and older, this difference persisted, with the highly educated group experiencing a significantly higher quality of life compared to the low educated group.

**Figure 3.**
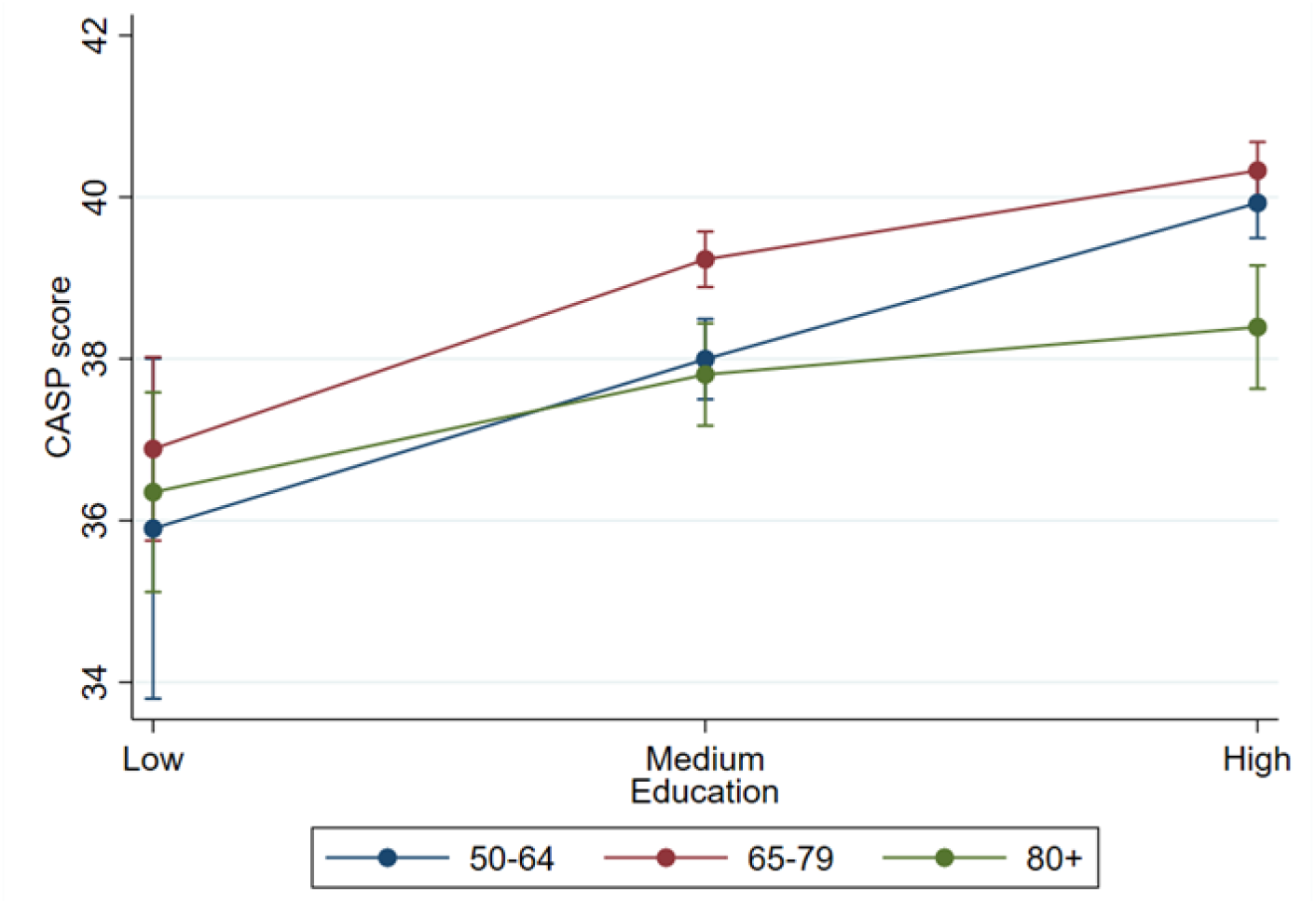
Quality of life by education and age. Data: SHARE Wave 9, Germany, weighted. N = 4,148.

### 3.4 Mediation Analysis

To examine the correlation between education and well-being, as well as the potential mediation of this correlation by self-rated health, health literacy and various health behaviours, a path analysis was conducted (the full model with all parameter estimates is presented in Appendix 3).

Controlling for various factors, including the financial situation of the respondents, revealed no significant direct effect of education on well-being. However, mediation analysis indicated only indirect effects for health literacy, fruit and vegetable consumption, and self-rated health (see Table 3). Specifically, the model showed significant correlations between education and health literacy, fruit and vegetable consumption, and health, and these three variables were significantly correlated with well-being. The mediation effect strongly contributed to the correlation between education and well-being. The indirect effect via self-rated health accounted for 54.7 percent of the total effect. The indirect effects via health literacy and fruit and vegetable consumption were 24.7 and 12.6 percent of the total effect, respectively. No mediating effect was found for any of the other tested health behaviours.

**Table 3:**
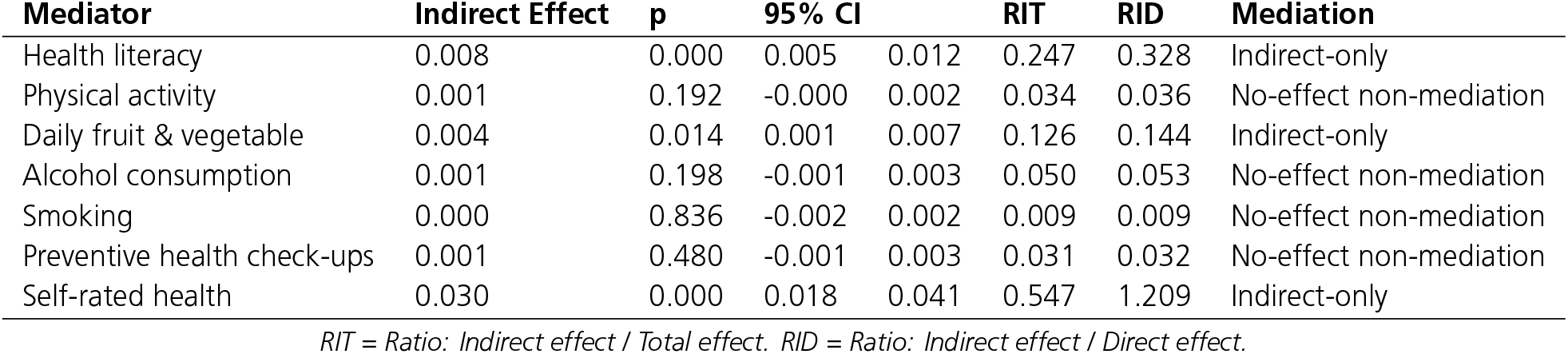
Indirect effects of education on well-being.

Health was the most important mediator between education and quality of life, but it was also potentially mediated by health behaviours themselves. The mediation of health literacy and health behaviours in the relationship between education and health was therefore tested (see table in Appendix 4). After accounting for all controls, education was found to have a direct positive association with self-rated health. Complementary mediation effects, i.e. direct and indirect effects in the same direction, were found for health literacy, physical activity, alcohol consumption, and smoking. Higher education was correlated with better health (direct effect) and with higher health literacy, which was also correlated with better health (indirect effect). Similarly, the indirect effects indicate that smoking was less common among the highly educated and associated with poorer health, while high physical activity was more common among the highly educated and correlated with better health. The indirect effect for alcohol consumption appeared counter-intuitive. Higher education was associated with a higher likelihood of alcohol consumption, while alcohol consumption was also correlated with better health.

The analysis revealed a competitive mediation for preventive health check-ups, i.e. the direct effect (higher education correlates with better health) is in the opposite direction to the indirect effect: The mediation analysis showed that higher education is connected to a higher number of preventive health check-ups, but more check-ups are connected to poorer health. Fruit and vegetable consumption showed “direct-only non-mediation”. Each of the complementary and competitive mediations contributed 3-7 percent of the total effect, except for health literacy, which contributed 8.9 percent.

Lastly, the direct effects of health literacy and health behaviours on well-being, as well as the potential mediation via self-rated health were tested (Appendix 5). All behaviours and health literacy were connected to well-being to some extent. Only indirect effects were observed for alcohol consumption, smoking, preventive health check-ups. Complementary effects were found for health literacy and physical activity. The indirect effect via fruit and vegetable consumption was slightly above significance (p=0.052) and therefore considered as indirect-only effect.

## 4 Discussion

This study analysed the correlations of education and well-being and the role of health literacy and health behaviours in this relationship. By analysing a sample of individuals age 50 years and older this study provides new insights into health behaviours and its influence on well-being of older adults in Germany. Well-being was measured using the CASP index of quality of life. The first research question addressed the associations of formal education with health literacy and preventive health behaviours. Education was significantly correlated with health literacy and all health behaviours (high physical activity, fruit and vegetable consumption, alcohol consumption, smoking, health check-ups). While most health behaviours tended to be more favourable in higher educated individuals, the opposite was found for alcohol consumption. This is consistent with previous findings of the general population in Germany (Lange et al., 2017; A. Richter et al., 2025). Age differences were evident in health literacy and physical activity, with the 80+ age group performing worse than younger age groups.

Second research question investigated if health literacy and health behaviours mediated the relationship between formal education and well-being in later life. Education correlated with well-being only via indirect path of mediators (self-rated health, health literacy, and fruit and vegetable consumption). Earlier studies found that financial situation mediates the correlation of education and well-being (Powdthavee et al., 2015; Sheikh et al., 2017). Although financial situation as a mediator was not tested in our analyses, indicators of the house-hold’s financial situation were included as control variables and financial situation correlated with both health and well-being. Including these variables as controls resulted in a non-significant correlation of education and well-being, which would be significant otherwise.

Only health literacy, high physical activity, and fruit and vegetable consumption had a direct effect on well-being. For health literacy and physical activity, the correlation with quality of life was additionally mediated via health. Other health behaviours showed only indirect effects on of quality of life mediated via health.

We found a higher number of check-ups to be correlated with worse health and therefore with worse well-being. Additional analyses revealed that this was driven by negative correlations of flu vaccination and eye examinations with health. This may be because eye examinations are mostly carried out when there are evident difficulties with eyesight (rather than as a preventive screening measure). Similarly, flu vaccinations may be recommended by doctors for vulnerable groups with poor health. Another unexpected finding was the correlation of better health with alcohol consumption. This might be caused by individuals with poor health not being able to drink alcohol at all, e.g., due to interactions between alcohol and medication.

Overall, health literacy and health behaviours correlated with well-being as mediators of the educational effect, as well as having a direct influence, and being mediated via health. Health literacy consistently had a stronger influence than single health behaviours. However, it should be noted that the Single Item Literacy Screener (SILS) was used as an indicator of health literacy. This specifically focuses on written health materials and does not represent a general measure of health literacy in its broadest sense. Another limitation is the cross-sectional nature of the data, which limits the ability to provide causal inference. While health behaviour was theoretically modelled to influence health, the opposite may be true, particularly with regard to physical activity, which may be limited due to health issues, and alcohol consumption, which may be reduced due to drug intake.

The results highlight the importance of health behaviours for well-being in old age. As these behaviours are directly or indirectly connected to well-being, and are modifiable, they are potential intervention targets. Although education was connected to all behaviours, only one behaviour, i.e. fruit and vegetable consumption, was found to mediate the relationship between education and well-being. These results suggest that interventions aimed at improving health behaviours may not necessarily need to be tailored according to educational level.

## Data Availability

All data produced are available online at the SHARE-ERIC website.

https://share-eric.eu/data/data-access

## Acknowledgements

The SHARE data collection has been funded by the European Commission, DG RTD through FP5 (QLK6-CT-2001-00360), FP6 (SHARE-I3: RII-CT-2006-062193, COMPARE: CIT5-CT-2005-028857, SHARELIFE: CIT4-CT-2006-028812), FP7 (SHARE-PREP: GA N°211909, SHARE-LEAP: GA N°227822, SHARE M4: GA N°261982, DASISH: GA N°283646) and Horizon 2020 (SHARE-DEV3: GA N°676536, SHARE-COHESION: GA N°870628, SERISS: GA N°654221, SSHOC: GA N°823782, SHARE-COVID19: GA N°101015924) and by DG Employment, Social Affairs & Inclusion through VS 2015/0195, VS 2016/0135, VS 2018/0285, VS 2019/0332, VS 2020/0313, SHARE-EUCOV: GA N°101052589 and EUCOVII: GA N°101102412. Additional funding from the German Federal Ministry of Education and Research (01UW1301, 01UW1801, 01UW2202), the Max Planck Society for the Advancement of Science, the U.S. National Institute on Aging (U01_AG09740-13S2, P01_AG005842, P01_AG08291, P30_AG12815, R21_AG025169, Y1-AG-4553-01, IAG_BSR06-11, OGHA_04-064, BSR12-04, R01_AG052527-02, R01_AG056329-02, R01_AG063944, HHSN271201300071C, RAG052527A) and from various national funding sources is gratefully acknowledged (see www.share-eric.eu).

This manuscript was produced in the context of the workshop ‘Empirical Analyses of Education and Learning in Later Life’, which was held in March 2026 at the Catholic University of Applied Sciences in Freiburg by the Expert Commission for the Tenth Government Report on Older People and the Office of the Government Reports on Older People, based at the German Centre of Gerontology (DZA). This workshop was made possible by funding from the Federal Ministry of Education, Family Affairs, Senior Citizens, Women and Youth (BMBFSFJ). We would like to thank Matthias Kliegel and Cornelia Kricheldorff for their helpful feedback on earlier versions of the manuscript.

This research project was conducted at the SHARE BERLIN Institute and funded by the German Federal Ministry of Research, Technology and Space (BMFTR) as part of SHARE (grant numbers 01UW1801 /01UW2202).

The SHARE data collection has been funded by the European Commission, DG RTD through FP5 (QLK6-CT-2001-00360), FP6 (SHARE-I3: RII-CT-2006-062193, COMPARE: CIT5-CT-2005-028857, SHARELIFE: CIT4-CT-2006-028812), FP7 (SHARE-PREP: GA N°211909, SHARE-LEAP: GA N°227822, SHARE M4: GA N°261982, DASISH: GA N°283646) and Horizon 2020 (SHARE-DEV3: GA N°676536, SHARE-COHESION: GA N°870628, SERISS: GA N°654221, SSHOC: GA N°823782, SHARE-COVID19: GA N°101015924) and by DG Employment, Social Affairs & Inclusion through VS 2015/0195, VS 2016/0135, VS 2018/0285, VS 2019/0332, VS 2020/0313, SHARE-EUCOV: GA N°101052589 and EUCOVII: GA N°101102412. Additional funding from the German Federal Ministry of Research, Technology and Space (01UW1301, 01UW1801, 01UW2202), Deutsche Forschungsge-meinschaft (549839254), the Max Planck Society for the Advancement of Science, the U.S. National Institute on Aging (U01_AG09740-13S2, P01_AG005842, P01_AG08291, P30_AG12815, R21_AG025169, Y1-AG-4553-01, IAG_BSR06-11, OGHA_04-064, BSR12-04, R01_AG052527-02, R01_AG056329-02, R01_AG063944, HHSN271201300071C, RAG052527A) and from various national funding sources is gratefully acknowledged (see www.share-eric.eu).

## A Appendix

### A.1 Health literacy and health behaviours by age group and education, weighted, without controls

| Age / education | High health literacy | High physical activity | Daily fruit & vegetables | Alcohol consumption | Smoking | Preventive Health check-ups |
| --- | --- | --- | --- | --- | --- | --- |
| 50-64 / Low | 0.735<br>[0.636,0.834] | 0.390<br>[0.277,0.503] | 0.546<br>[0.422,0.670] | 0.411<br>[0.289,0.533] | 0.442<br>[0.318,0.567] | 1.966<br>[1.756,2.175] |
| 50-64 / Medium | 0.861<br>[0.835,0.887] | 0.571<br>[0.531,0.610] | 0.661<br>[0.622,0.699] | 0.603<br>[0.563,0.643] | 0.304<br>[0.265,0.343] | 2.069<br>[1.985,2.154] |
| 50-64 / High | 0.928<br>[0.908,0.949] | 0.518<br>[0.476,0.561] | 0.674<br>[0.633,0.714] | 0.706<br>[0.667,0.746] | 0.192<br>[0.155,0.229] | 2.256<br>[2.169,2.343] |
| 65-79 / Low | 0.751<br>[0.678,0.824] | 0.234<br>[0.163,0.305] | 0.713<br>[0.635,0.791] | 0.459<br>[0.374,0.545] | 0.183<br>[0.115,0.250] | 2.104<br>[1.912,2.296] |
| 65-79 / Medium | 0.860<br>[0.837,0.882] | 0.339<br>[0.307,0.371] | 0.752<br>[0.723,0.781] | 0.578<br>[0.545,0.610] | 0.152<br>[0.128,0.177] | 2.412<br>[2.340,2.483] |
| 65-79 / High | 0.888<br>[0.866,0.911] | 0.453<br>[0.417,0.490] | 0.789<br>[0.759,0.820] | 0.702<br>[0.669,0.736] | 0.110<br>[0.0865,0.133] | 2.559<br>[2.485,2.632] |
| 80+ / Low | 0.599<br>[0.487,0.712] | 0.120<br>[0.0358,0.205] | 0.720<br>[0.618,0.823] | 0.360<br>[0.252,0.468] | 0.0592<br>[0.00213,0.116] | 2.307<br>[2.077,2.537] |
| 80+ / Medium | 0.662<br>[0.605,0.719] | 0.205<br>[0.157,0.252] | 0.784<br>[0.734,0.834] | 0.503<br>[0.443,0.564] | 0.0376<br>[0.0151,0.0602] | 2.317<br>[2.195,2.439] |
| 80+ / High | 0.742<br>[0.676,0.809] | 0.269<br>[0.203,0.334] | 0.870<br>[0.822,0.918] | 0.582<br>[0.509,0.655] | 0.0529<br>[0.0181,0.0877] | 2.485<br>[2.331,2.638] |
| N | 4148 | 4148 | 4148 | 4148 | 4148 | 4148 |

Data: SHARE wave 9, Germany, 95% confidence intervals in brackets.

### A.2 CASP by education and age

| Education | Age |  |  |
| --- | --- | --- | --- |
|  | 50-64 | 65-79 | 80+ |
| Low | 35.90<br>[33.80,38.00] | 36.89<br>[35.75,38.02] | 36.35<br>[35.12,37.58] |
| Medium | 38.00<br>[37.50,38.49] | 39.23<br>[38.89,39.57] | 37.81<br>[37.17,38.44] |
| High | 39.93<br>[39.49,40.36] | 40.33<br>[39.97,40.68] | 38.39<br>[37.63,39.16] |
| Observations | 4,148 |  |  |

Data: SHARE wave 9, Germany, 95% confidence intervals in brackets.

### A.3 Path analysis (Linear regressions)

|  | **Coef.** | **Std. Err.** | **p** | **95% CI** |  |
| --- | --- | --- | --- | --- | --- |
| Self-rated health | 2.06 | 0.08 | 0.000 | 1.90 | 2.21 |
| Health literacy | 1.39 | 0.20 | 0.000 | 1.00 | 1.78 |
| High physical activity | 0.29 | 0.15 | 0.049 | 0.00 | 0.59 |
| Daily fruit & vegetable | 0.46 | 0.17 | 0.005 | 0.14 | 0.79 |
| Alcohol consumption | 0.21 | 0.15 | 0.158 | -0.08 | 0.51 |
| Smoking | -0.04 | 0.20 | 0.835 | -0.43 | 0.35 |
| Preventive health check-ups | 0.05 | 0.07 | 0.459 | -0.08 | 0.19 |
| Education | 0.22 | 0.12 | 0.083 | -0.03 | 0.46 |
| Age: 65–79 | 0.91 | 0.16 | 0.000 | 0.60 | 1.23 |
| Age: 80+ | 0.56 | 0.25 | 0.024 | 0.08 | 1.05 |
| Gender | 0.28 | 0.15 | 0.066 | -0.02 | 0.58 |
| Partner in household | 0.69 | 0.17 | 0.000 | 0.36 | 1.01 |
| 10-word memory test | 0.17 | 0.05 | 0.000 | 0.07 | 0.26 |
| Born outside Germany | -0.52 | 0.23 | 0.027 | -0.98 | -0.06 |
| Good eyesight: reading | 0.72 | 0.24 | 0.002 | 0.25 | 1.19 |
| Make ends meet: great difficulties | -5.20 | 0.47 | 0.000 | -6.12 | -4.28 |
| Make ends meet: some difficulties | -2.55 | 0.27 | 0.000 | -3.09 | -2.01 |
| Poverty risk: Yes | -1.05 | 0.23 | 0.000 | -1.51 | -0.60 |
| _cons | 29.02 | 0.48 | 0.000 | 28.07 | 29.97 |

*Path analysis: Dependent variable – Self-rated health*
|  | **Coef.** | **Std. Err.** | **p** | **95% CI** |  |
| --- | --- | --- | --- | --- | --- |
| High reading health literacy | 0.24 | 0.04 | 0.000 | 0.16 | 0.32 |
| High physical activity | 0.29 | 0.03 | 0.000 | 0.23 | 0.35 |
| Daily fruit & vegetable | 0.06 | 0.03 | 0.050 | 0.00 | 0.13 |
| Alcohol consumption | 0.19 | 0.03 | 0.000 | 0.13 | 0.24 |
| Smoking | -0.15 | 0.04 | 0.000 | -0.23 | -0.07 |
| Preventive health check-ups | -0.03 | 0.01 | 0.025 | -0.06 | 0.00 |
| Education | 0.13 | 0.02 | 0.000 | 0.08 | 0.17 |
| Age: 65–79 | -0.10 | 0.03 | 0.001 | -0.16 | -0.04 |
| Age: 80+ | -0.26 | 0.05 | 0.000 | -0.35 | -0.16 |
| Gender | -0.06 | 0.03 | 0.058 | -0.12 | 0.00 |
| Partner in household | 0.00 | 0.03 | 0.997 | -0.07 | 0.06 |
| 10-word memory test | 0.08 | 0.01 | 0.000 | 0.06 | 0.10 |
| Born outside Germany | -0.01 | 0.05 | 0.876 | -0.10 | 0.08 |
| Good eyesight: reading | 0.23 | 0.05 | 0.000 | 0.14 | 0.32 |
| Make ends meet: great difficulties | -0.56 | 0.09 | 0.000 | -0.75 | -0.38 |
| Make ends meet: some difficulties | -0.32 | 0.05 | 0.000 | -0.43 | -0.22 |
| Poverty risk: Yes | -0.06 | 0.05 | 0.183 | -0.15 | 0.03 |
| _cons | 1.80 | 0.09 | 0.000 | 1.62 | 1.98 |

*Path analysis: Dependent variable – Health literacy*
|  | **Coef.** | **Std. Err.** | **p** | **95% CI** |  |
| --- | --- | --- | --- | --- | --- |
| Education | 0.05 | 0.01 | 0.000 | 0.03 | 0.07 |
| Age: 65–79 | 0.00 | 0.01 | 0.700 | -0.02 | 0.03 |
| Age: 80+ | -0.13 | 0.02 | 0.000 | -0.16 | -0.09 |
| Gender | -0.01 | 0.01 | 0.421 | -0.03 | 0.01 |
| Partner in household | -0.03 | 0.01 | 0.016 | -0.06 | -0.01 |
| 10-word memory test | 0.03 | 0.00 | 0.000 | 0.02 | 0.03 |
| Born outside Germany | -0.03 | 0.02 | 0.166 | -0.06 | 0.01 |
| Good eyesight: reading | 0.09 | 0.02 | 0.000 | 0.06 | 0.13 |
| Make ends meet: great difficulties | -0.07 | 0.04 | 0.054 | -0.14 | 0.00 |
| Make ends meet: some difficulties | 0.01 | 0.02 | 0.795 | -0.04 | 0.05 |
| Poverty risk: Yes | -0.02 | 0.02 | 0.257 | -0.06 | 0.01 |
| _cons | 0.61 | 0.03 | 0.000 | 0.55 | 0.67 |

*Path analysis: Dependent variable – Physical activity*
|  | **Coef.** | **Std. Err.** | **p** | **95% CI** |  |
| --- | --- | --- | --- | --- | --- |
| Education | 0.03 | 0.01 | 0.043 | 0.00 | 0.05 |
| Age: 65–79 | -0.12 | 0.02 | 0.000 | -0.15 | -0.08 |
| Age: 80+ | -0.23 | 0.03 | 0.000 | -0.28 | -0.18 |
| Gender | 0.04 | 0.02 | 0.008 | 0.01 | 0.07 |
| Partner in household | 0.02 | 0.02 | 0.192 | -0.01 | 0.06 |
| 10-word memory test | 0.03 | 0.00 | 0.000 | 0.02 | 0.04 |
| Born outside Germany | -0.06 | 0.02 | 0.016 | -0.11 | -0.01 |
| Good eyesight: reading | 0.05 | 0.03 | 0.030 | 0.01 | 0.10 |
| Make ends meet: great difficulties | 0.03 | 0.05 | 0.540 | -0.07 | 0.13 |
| Make ends meet: some difficulties | 0.02 | 0.03 | 0.509 | -0.04 | 0.08 |
| Poverty risk: Yes | -0.09 | 0.02 | 0.000 | -0.14 | -0.04 |
| _cons | 0.25 | 0.04 | 0.000 | 0.17 | 0.34 |

*Path analysis: Dependent variable – Fruit & vegetable consumption*
|  | **Coef.** | **Std. Err.** | **p** | **95% CI** |  |
| --- | --- | --- | --- | --- | --- |
| Education | 0.07 | 0.01 | 0.000 | 0.04 | 0.09 |
| Age: 65–79 | 0.10 | 0.01 | 0.000 | 0.07 | 0.13 |
| Age: 80+ | 0.16 | 0.02 | 0.000 | 0.11 | 0.20 |
| Gender | -0.19 | 0.01 | 0.000 | -0.21 | -0.16 |
| Partner in household | 0.06 | 0.02 | 0.000 | 0.03 | 0.09 |
| 10-word memory test | 0.00 | 0.00 | 0.428 | -0.01 | 0.01 |
| Born outside Germany | 0.05 | 0.02 | 0.036 | 0.00 | 0.09 |
| Good eyesight: reading | 0.01 | 0.02 | 0.785 | -0.04 | 0.05 |
| Make ends meet: great difficulties | -0.10 | 0.04 | 0.018 | -0.19 | -0.02 |
| Make ends meet: some difficulties | 0.02 | 0.03 | 0.515 | -0.03 | 0.07 |
| Poverty risk: Yes | -0.04 | 0.02 | 0.087 | -0.08 | 0.01 |
| _cons | 0.64 | 0.04 | 0.000 | 0.57 | 0.71 |

*Path analysis: Dependent variable – Alcohol consumption*
|  | **Coef.** | **Std. Err.** | **p** | **95% CI** |  |
| --- | --- | --- | --- | --- | --- |
| Education | 0.05 | 0.01 | 0.000 | 0.03 | 0.08 |
| Age: 65–79 | -0.01 | 0.02 | 0.724 | -0.04 | 0.03 |
| Age: 80+ | -0.04 | 0.02 | 0.073 | -0.09 | 0.00 |
| Gender | 0.13 | 0.02 | 0.000 | 0.10 | 0.16 |
| Partner in household | 0.07 | 0.02 | 0.000 | 0.03 | 0.10 |
| 10-word memory test | 0.02 | 0.00 | 0.000 | 0.01 | 0.03 |
| Born outside Germany | -0.10 | 0.02 | 0.000 | -0.14 | -0.05 |
| Good eyesight: reading | 0.03 | 0.02 | 0.269 | -0.02 | 0.08 |
| Make ends meet: great difficulties | -0.07 | 0.05 | 0.121 | -0.17 | 0.02 |
| Make ends meet: some difficulties | -0.04 | 0.03 | 0.122 | -0.10 | 0.01 |
| Poverty risk: Yes | -0.15 | 0.02 | 0.000 | -0.20 | -0.10 |
| _cons | 0.34 | 0.04 | 0.000 | 0.26 | 0.42 |

*Path analysis: Dependent variable – Smoking*
|  | **Coef.** | **Std. Err.** | **p** | **95% CI** |  |
| --- | --- | --- | --- | --- | --- |
| Education | -0.05 | 0.01 | 0.000 | -0.07 | -0.03 |
| Age: 65–79 | -0.12 | 0.01 | 0.000 | -0.14 | -0.09 |
| Age: 80+ | -0.23 | 0.02 | 0.000 | -0.27 | -0.20 |
| Gender | 0.04 | 0.01 | 0.001 | 0.02 | 0.06 |
| Partner in household | -0.08 | 0.01 | 0.000 | -0.10 | -0.05 |
| 10-word memory test | -0.01 | 0.00 | 0.026 | -0.02 | 0.00 |
| Born outside Germany | -0.04 | 0.02 | 0.036 | -0.07 | 0.00 |
| Good eyesight: reading | -0.05 | 0.02 | 0.012 | -0.08 | -0.01 |
| Make ends meet: great difficulties | 0.14 | 0.04 | 0.000 | 0.07 | 0.22 |
| Make ends meet: some difficulties | 0.03 | 0.02 | 0.131 | -0.01 | 0.07 |
| Poverty risk: Yes | 0.07 | 0.02 | 0.000 | 0.03 | 0.10 |
| _cons | 0.42 | 0.03 | 0.000 | 0.36 | 0.48 |

*Path analysis: Dependent variable – Preventive check-ups*
|  | **Coef.** | **Std. Err.** | **p** | **95% CI** |  |
| --- | --- | --- | --- | --- | --- |
| Education | 0.14 | 0.03 | 0.000 | 0.08 | 0.19 |
| Age: 65–79 | 0.34 | 0.04 | 0.000 | 0.27 | 0.41 |
| Age: 80+ | 0.35 | 0.05 | 0.000 | 0.24 | 0.45 |
| Gender | -0.19 | 0.03 | 0.000 | -0.26 | -0.13 |
| Partner in household | 0.21 | 0.04 | 0.000 | 0.13 | 0.28 |
| 10-word memory test | 0.02 | 0.01 | 0.045 | 0.00 | 0.04 |
| Born outside Germany | -0.17 | 0.05 | 0.002 | -0.27 | -0.06 |
| Good eyesight: reading | -0.02 | 0.05 | 0.662 | -0.13 | 0.08 |
| Make ends meet: great difficulties | -0.19 | 0.11 | 0.078 | -0.40 | 0.02 |
| Make ends meet: some difficulties | -0.02 | 0.06 | 0.783 | -0.14 | 0.11 |
| Poverty risk: Yes | -0.32 | 0.05 | 0.000 | -0.42 | -0.21 |
| _cons | 1.87 | 0.09 | 0.000 | 1.70 | 2.05 |

LR test of model vs. saturated: $\chi^2(15) = 152.85$ , $Prob > \chi^2 = 0.0000$ . Data: SHARE wave 9, Germany.
Data: SHARE wave 9, Germany.

### A.4 Indirect effects of education on self-reported health

| Mediator | Indirect Effect | p-value | 95% CI | RIT | RID | Mediation |
| --- | --- | --- | --- | --- | --- | --- |
| Health literacy | 0.008 | 0.000 | 0.004 , 0.012 | 0.089 | 0.097 | Complementary |
| Physical activity | 0.005 | 0.049 | 0.000 , 0.010 | 0.057 | 0.060 | Complementary |
| Daily fruit & vegetable | 0.003 | 0.069 | -0.000 , 0.006 | 0.033 | 0.034 | Direct-only non-mediation |
| Alcohol consumption | 0.006 | 0.001 | 0.003 , 0.010 | 0.073 | 0.078 | Complementary |
| Smoking | 0.004 | 0.003 | 0.002 , 0.008 | 0.051 | 0.054 | Complementary |
| Preventive health check-ups | -0.003 | 0.047 | -0.005 , -0.000 | 0.034 | 0.032 | Competitive |

RIT= Ratio: Indirect effect / Total effect. RID = Ratio: Indirect effect / Direct effect. Data: SHARE wave 9, Germany.

### A.5 Direct and indirect effects of health literacy and health behaviours on well-being, mediated via self-rated health

| Independent Variable | Indirect Effect | p | 95% CI | RIT | RID | Direct effect | Mediation |
| --- | --- | --- | --- | --- | --- | --- | --- |
| Health literacy | 0.033 | 0.000 | 0.022, 0.044 | 0.264 | 0.358 | positive | Complementary |
| Physical activity | 0.054 | 0.000 | 0.043, 0.066 | 0.671 | 2.044 | positive | Complementary |
| Daily fruit & vegetable | 0.011 | 0.052 | -0.000, 0.021 | 0.223 | 0.287 | positive | Direct-only |
| Alcohol consumption | 0.034 | 0.000 | 0.023, 0.045 | 0.641 | 1.788 | non-sig. | Indirect-only |
| Current smoker | -0.021 | 0.000 | -0.032, -0.010 | 0.879 | 7.274 | non-sig. | Indirect-only |
| Preventive health check-ups | -0.012 | 0.025 | -0.023, -0.002 | 5.326 | 1.231 | non-sig. | Indirect-only |

RIT= Ratio: Indirect effect / Total effect. RID = Ratio: Indirect effect / Direct effect. Data: SHARE wave 9, Germany.

## Notes

### Competing Interest Statement

The authors have declared no competing interest.

### Author Declarations

The SHARE data are distributed by SHARE-ERIC (Survey of Health, Ageing and Retirement in Europe) to registered users through the SHARE Research Data Center. Access to the data collected and generated in the SHARE projects is provided free of charge for scientific use globally. The dataset used in our study (SHARE) were de-identified prior to use. The data are provided as scientific-use files and comply with strict data protection and privacy regulations. In particular, SHARE data are released under conditions of factual anonymity.

